# Colorectal Cancer Disparities Across the Continuum of Cancer Care: A Systematic Review and Meta-Analysis

**DOI:** 10.1101/2021.07.01.21259880

**Authors:** Solomiya Syvyk, Sanford E. Roberts, Caitlin B. Finn, Chris Wirtalla, Rachel Kelz

## Abstract

**Background and Objectives:** Disparate colorectal cancer outcomes persist in vulnerable populations. We aimed to examine the distribution of research across the colorectal cancer care continuum, and to determine disparities in the use of Surgery among Black patients.

**Methods:** A systematic review and meta-analysis of colorectal cancer disparities studies was performed. The meta-analysis assessed three utilization measures in Surgery.

**Results:** Of 1,199 publications, 60% focused on Prevention, Screening, or Diagnosis, 20% on Survivorship, 15% on Treatment, and 1% on End-of-Life Care. A total of 16 studies, including 1,110,674 patients, were applied to three separate meta-analyses regarding utilization of Surgery. Black colorectal cancer patients were less likely to receive surgery, twice as likely to refuse surgery, and less likely to receive laparoscopic surgery when compared to White patients.

**Conclusions:** Over the past 10 years, the majority of published research remained focused on the prevention, screening, or diagnosis domain. Given the observed treatment disparities and persistently elevated disease-specific mortality among Black patients, future efforts to reduce colorectal cancer disparities should include interventions within Surgery.

**Synopsis:** In this systematic review on disparities along the colorectal cancer care continuum, we found that 64% of research has been focused on prevention, screening, or diagnosis while only 6% addressed surgical disparities. In the meta-analysis, Black patients were less likely to undergo surgery, more likely to refuse surgery, and less likely to undergo laparoscopic surgery, when compared to White patients. Future research should target treatment differences across populations in order to impact persistent disparities in colorectal cancer survival.

## Introduction

Despite substantial progress in care for patients with colorectal cancer over the past several decades, these advances have been unevenly distributed. While colorectal cancer mortality rates have decreased for Black and White patients at all stages, the declines have been smaller for Black patients by 15%-28% at every stage.^1^ Similarly, the Black community continues to suffer from worse adverse colorectal cancer outcomes such as higher risk-adjusted post-operative complications and lower overall survival rates. ^2^ The mortality difference may reflect lower utilization of recommended colorectal cancer treatment among Black patients when compared to White patients.^3^

The majority of research and policy work in colorectal cancer disparities performed before 2010 was focused on prevention, screening and diagnosis.^4,5^ Over two decades, there were more than 230 publications and numerous policy and clinical efforts focused on interventions to increase colorectal cancer screening for Black patients. These efforts were associated with an increase in colorectal cancer screening rates from 32% (2000) to 59% (2016) among Black patients which was similar to rates among White patients.^6,7^

Effective efforts to ameliorate disparities in colorectal cancer outcomes have been focused on prevention, screening, and diagnosis. Little is known about efforts to eradicate disparities across the remainder of the colorectal cancer care continuum. We conducted this systematic review and meta-analysis to guide future research efforts. To that end, first, we assessed the volume of colorectal cancer disparities publications attributed to each domain of the cancer care continuum. Secondly, because Surgery is the primary and most common treatment for non-metastatic colorectal cancer, we examined disparities within the utilization of Surgery, as a treatment modality.

## Materials and Methods

### Protocol

We performed this systemic review and meta-analysis in accordance with the Preferred Reporting Items for Systematic Reviews and Meta-Analyses (PRISMA) guidelines.^8^ A preliminary version was published on medRxiv. Since its posting, the study has been significantly modified.

### Information Sources and Search Strategies

We performed a systematic literature search of MEDLINE and Scopus databases using search terms related to colorectal cancer disparities. Table 1 in the Supplement gives the search strategy used for MEDLINE. Within the Scopus database, the following search terms were applied: “colorectal” AND “disparities”. Our searches were limited to studies with full manuscripts, published in English, and from January 1, 2011 to March 29, 2021.

**Table I.** Distribution of Treatment Disparities Studies within Colorectal Cancer by Treatment Type and Study Population.

| Distribution of Treatment Disparities Studies within Colorectal Cancer by Treatment Type |  |  |  |  |  |  |
| --- | --- | --- | --- | --- | --- | --- |
| Treatment Type | Surgery | Combined |  | Radiation |  | Systemic Therapy |
|  |  | w/ Surgery | w/o Surgery |  |  |  |
| Publications, n (%) | 69 (6%) | 26 (2%) | 37 (3%) | 13 (1%) |  | 33 (3%) |
| Distribution of Treatment Disparities Studies within Colorectal Cancer by Study Population |  |  |  |  |  |  |
| Disparate Population | Race and Ethnicity | SES | Age | Gender | Disabilities/Comorbidities | LGBTQI+ |
| Publications, n (%) | 82 (46%) | 50 (28%) | 30 (17%) | 10 (6%) | 6 (3%) | 0 (0%) |
| Note. Percentages for the type of treatment are calculated out of the total number of disparities studies; Percentages for the study population are calculated out of the total number of publications on treatment disparities |  |  |  |  |  |  |

### Eligibility Criteria

We included studies that reported on colon, rectal, or colorectal disparities within the United States. We considered studies along the entire cancer care continuum, as per the Institute of Medicine Framework.^9^ Commentaries, letters, and publications reporting on populations outside of the United States were excluded. Review articles were excluded from the primary analysis, but their reference lists were used as a source of additional relevant articles.

The meta-analysis was designed specifically to examine disparities in the utilization of Surgery as a treatment modality. For the meta-analysis, we included studies relating to disparities in the utilization of Surgery: receipt of surgery, refusal of surgery, and receipt of laparoscopic versus open surgery. In accordance with the results of the systematic review, we tailored our meta-analyses to patients of Black race.

### Study Selection and Data Collection

Two authors (S.R. and S.S.) independently screened study titles and abstracts for potential inclusion. Full text of the relevant studies was extracted and reviewed for eligibility. Disagreements between the reviewers were resolved by consensus or by the third author (R.R.K). We manually searched reference lists of review articles for pertinent additional studies. Two authors (S.R. and S.S.) independently extracted data from the included studies and discrepancies were resolved by discussion.

### Data Items

We captured the following: (1) General Study Information including: title, author(s), year of publication, data source, and sample size; (2) Primary Disparity including: race and ethnicity, SES (income level, insurance status, location, hospital effects, or education level), age, gender, comorbidities/disabilities, and LGBTQIA+; (3) Categorization by the Cancer Care Continuum including: Prevention, Screening, or Diagnosis, Treatment, Survivorship, or End-of-Life Care; (4) Treatment Type including: Radiation, Systemic Therapy, Surgery, or Combined Treatment; (5) Utilization of Surgery Outcome(s) including: receipt of surgery, refusal of surgery, and receipt of laparoscopic versus open surgery. For studies included in the meta-analyses, we extracted the odds ratios (ORs), corresponding confidence intervals (CIs), the focus and control group sample sizes, and the covariates.

### Risk of Bias in Individual Studies

Two authors (S.R. and S.S.) independently assessed the studies included in the meta-analysis for potential bias using the Newcastle-Ottawa Scale.^10^ This scale assesses the potential of bias in 3 domains: (1) selection of the study groups; (2) comparability of groups; and (3) ascertainment of exposure and outcome. The maximum score in the selection domain is 4 stars, in the comparability domain is 2 stars, and in the outcome domain is 3 stars. Studies with scores of 7 or higher were considered as having a low risk of bias, scores of 4 to 6 as having a moderate risk of bias, and scores less than 4 as having a high risk of bias. Disagreements were resolved by consensus or by the third author (R.R.K).

### Statistical Analysis

We calculated combined estimates for the five separate analyses focused on utilization of Surgery outcomes: receipt of surgery (colorectal, colon, and rectal), refusal of surgery, and receipt of laparoscopic versus open surgery. First, we studied the association between receipt of surgery and Black race using ORs and corresponding 95% CIs from the multivariate analyses presented in the included studies. Given that some studies reported their results for colorectal procedures combined while others reported colon and rectal separately, we conducted three separate analyses for each procedure type: colorectal, colon, and rectal.

Next, we calculated pooled multivariate ORs and the associated 95% CIs for the association between refusal of colon surgery and Black race. In the fifth and final analysis, we calculated the association between receipt of laparoscopic versus open surgery and Black race; multivariate ORs and corresponding 95% CIs were obtained from the relevant studies.

When data was unclear or did not provide the appropriate outcome, the study was not included in the analysis for the outcome. A funnel plot and regression asymmetry test were originally planned to assess for small study bias, but could not be performed due the limited number of studies in each analysis.

The Cochran’s test was used to assess for heterogeneity of the included studies in each respective analysis. For the heterogeneity measure, derived from two-tailed tests, p values less than 0.10 were deemed to indicate significance. Forest plots and the *I*^2^ statistic results were also assessed with *I*^2^ > 50% indicating moderate heterogeneity. If heterogeneity was observed (p⍰<⍰.10 or I^2^⍰>⍰50%), a random-effects model was used to pool the estimate across studies, per analysis, with the DerSimonian-Laird method. Otherwise, a fixed-effects model was applied.

All database search results were downloaded, merged, and deduplicated by the systematic review management software, Covidence (Veritas Health Innovation, Melbourne, Australia). All analyses were performed using Stata statistical software version 16.1 (StataCorp, College Station, TX).

## Results

A total of 2,674 potentially relevant publications were title and abstract reviewed (Figure 1). Among them, 1,199 met the inclusion criteria for this systematic review. Over 60% of all included publications were focused on Prevention, Screening, or Diagnosis, followed by Survivorship (20%), Treatment (15%), and End-of-Life Care (1%) (Figure 2).

**Figure I.**
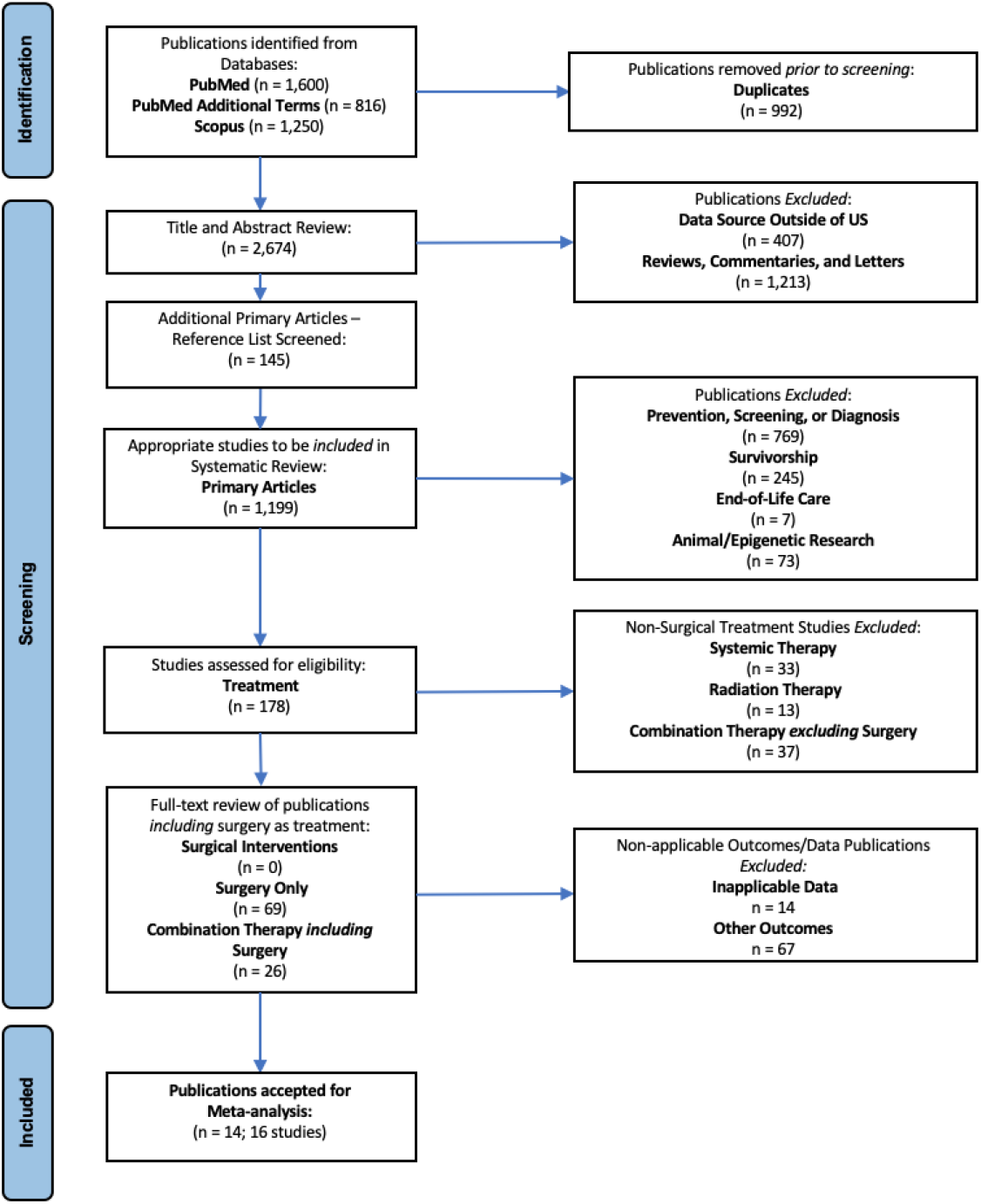
Flow Diagram of Study Disposition.

**Figure 2.**
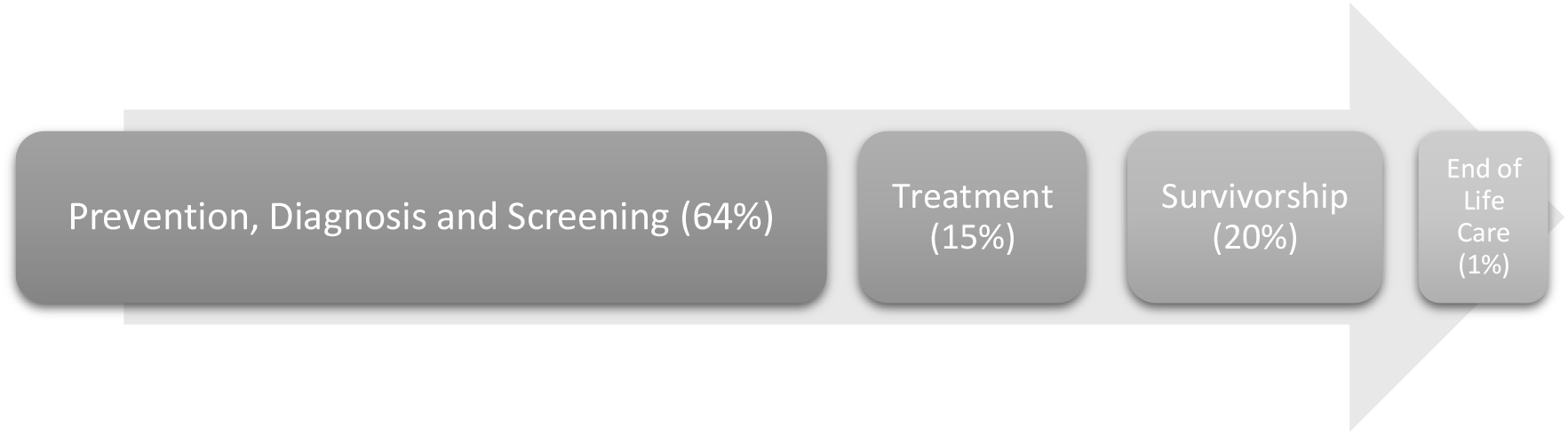
Proportion of Studies Published on Disparities in Colorectal Cancer Care Across the Cancer Care Continuum.

Within the studies reporting on treatment disparities, 46% were focused on Race and Ethnicity, followed by SES (28%), Age (17%), Gender (6%), Disabilities/Comorbidities (3%), and LGBTQI+ (0.0%). Within the treatment category, the most commonly examined disparate population was Black race. Of 95 articles that included surgical data, 69/1,199 (6%) focused exclusively on surgical disparities and 26/1,199 (2%) included Surgery as part of a combined treatment (Table 1).

Sixty-seven of the total 95 surgery-related articles reported on other outcomes such as pain management or delay in treatment and fourteen either did not include multivariate analysis, odds ratios, or 95% CIs. Ultimately, 14 publications (16 studies) were included in the meta-analysis as three separate analyses: receipt of colorectal cancer surgery, refusal of colon surgery, and receipt of laparoscopic versus open surgery and patients of Black race.^11-24^ Bliton et. al and Samuel et. al were applied to both receipt of colorectal cancer (colon) surgery and Black race and receipt of colorectal cancer (rectal) surgery and Black race without patient overlap. All 14 included publications were retrospective, and the total number of patients included was 1,110,674 (Table 2 in the Supplement).

### Risk of Bias Within Studies

The quality of studies ranged between 6 and 7, indicating predominantly high quality with low risk of bias. For comparability of groups, the variables chosen were a measure for patient comorbidities and stage of cancer. All of the studies reported a loss-to-follow up due to their retrospective nature and inherent limitations within certain national databases. (Table 3 in the Supplement).

### Meta-analyses

#### Receipt of Surgery and Black Race

A total of five studies evaluated the receipt of colorectal cancer surgery with moderate heterogeneity (*I*^2^ = 94.3%; p = 0.00). In the random-effects model used to obtain pooled results, Black patients with colorectal cancer were less likely to undergo Surgery (OR 0.75, 95% CI 0.60-0.93) when compared to White patients.

The additional two analyses by cancer type, colon or rectal, produced similar results, but with low between-study heterogeneity, (colon: *I*^2^ = 0.0%; p = 0.955) and (rectal: *I*^2^ = 39.2%; p = 0.193). The analyses by colon or rectal cancer included an additional six studies that were specific to the cancer type: colon (n=3) and rectal (n=3). Black race was associated with a lower likelihood of receiving colon cancer surgery (OR 0.78, 95% CI 0.74-0.83); and a lower likelihood of receiving rectal cancer surgery (OR 0.73, 95% CI 0.65-0.81) (Figure 3).

**Figure 3.**
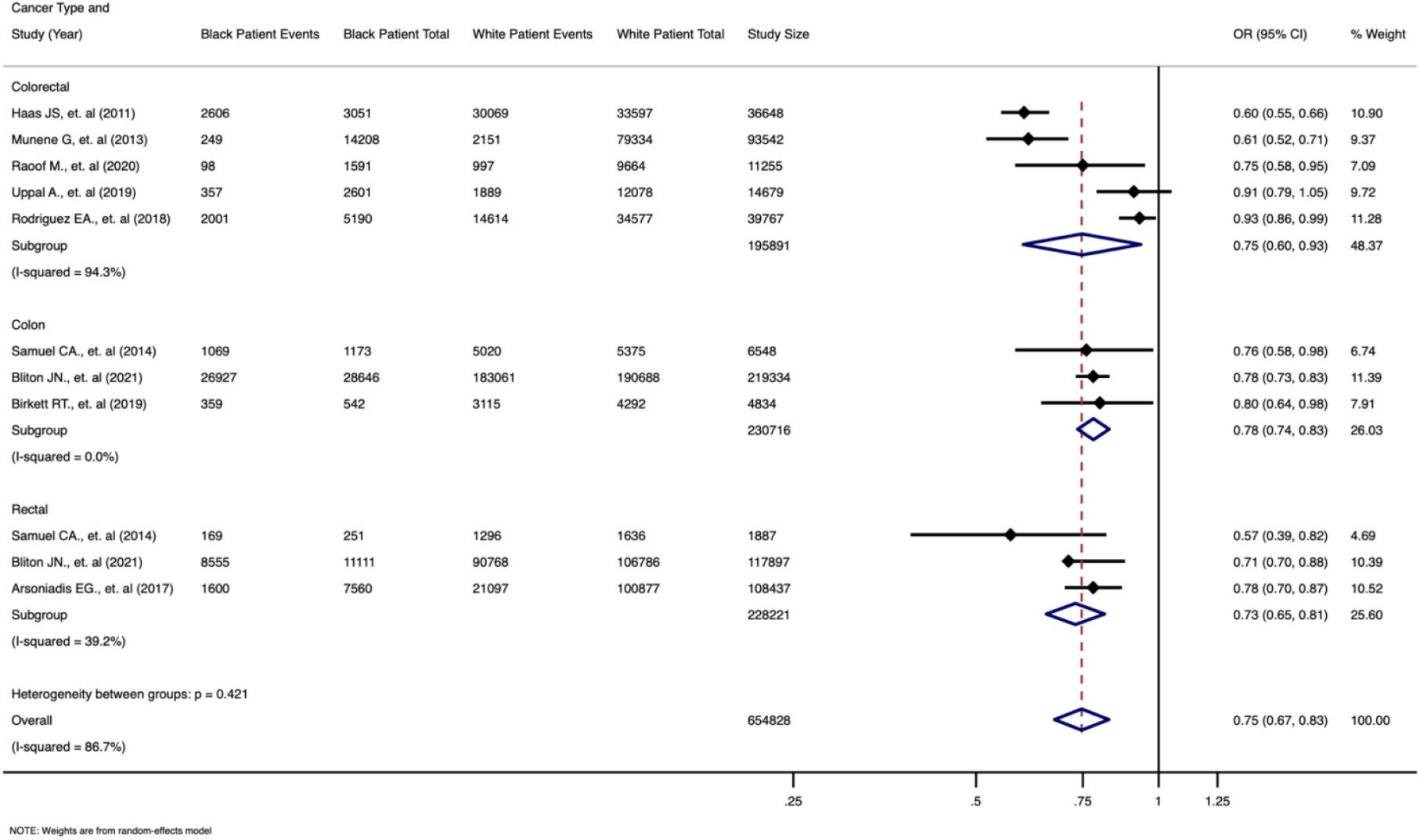
Meta-analysis of 9 Studies Assessing Receipt of Colorectal, Colon, or Rectal Cancer Surgery and Black Race.

#### Refusal of Colon Surgery and Black Race

The 2 studies on refusal of colon surgery and Black race demonstrated moderate heterogeneity (I^2^ = 61.7%; p = 0.106). Applying the random-effects model, our pooled analysis indicated that Black patients are more likely to refuse colon surgery than White patients (OR 2.41, 95% CI 1.91-3.06) (Figure 4).

**Figure 4.**
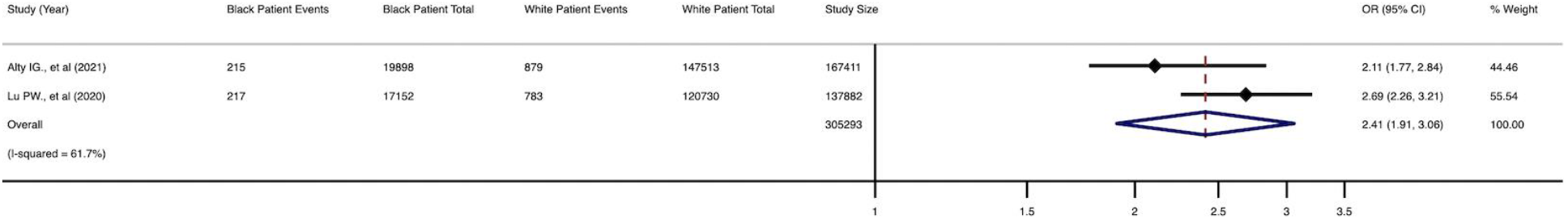
Meta-analysis of 2 Studies Assessing Refusal of Colon Cancer Surgery and Black Race.

#### Receipt of Laparoscopic versus Open colorectal cancer Surgery and Black Race

There was limited between-study heterogeneity presented in the three publications which assessed the receipt of laparoscopic versus open colorectal cancer surgery and Black race (I^2^ = 0.0%; p = 0.977). In our pooled analysis, Black patients were less likely to receive laparoscopic versus open colorectal cancer surgery when compared to White patients (OR 0.91, 95% CI 0.88-0.94) (Figure 5).

**Figure 5.**
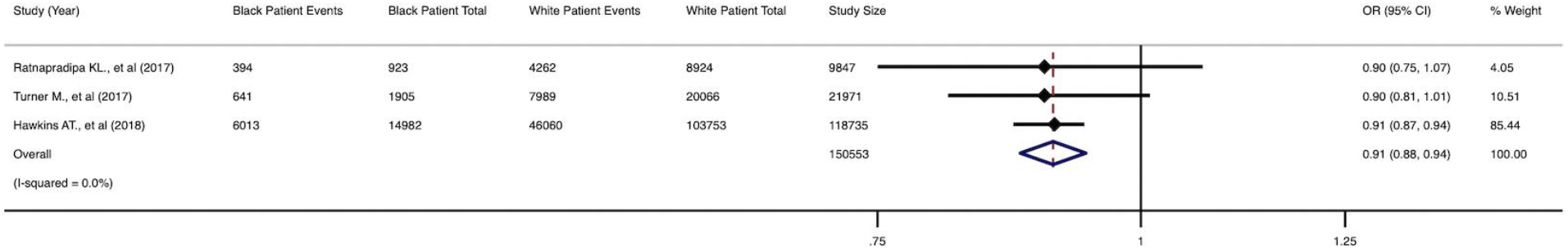
Meta-analysis of 3 Studies Assessing Receipt of Laparoscopic versus Open Colorectal Cancer Surgery and Black Race.

## Discussion

In 2002, the Institute of Medicine’s report titled “Unequal Treatment: Confronting Racial and Ethnic Disparities in Health Care”, summarized the multi-factorial roots of racial disparities, including patient, provider, and systemic factors.^25^ Since the publication of that report, the majority of the published research has remained focused on prevention, screening and diagnosis. Our approach offers researchers perspective into what areas within colorectal cancer disparities research may be saturated versus those which remain unexplored or underexplored.

We identified a disproportionate research focus on prevention, screening, or diagnosis at 64% of all included studies – four times the volume of studies focused on treatment (15%) and a tenfold difference to publications focused on Surgery as a treatment (6%). Within the treatment studies, the majority focused on racial minorities (46%) and low socioeconomic status (28%) and few addressed disparities among the LGBTQI+ population. Out of the 1,199 primary articles, none of the studies examined interventions designed to address disparities in Surgery.

Our findings are consistent with previous literature, citing the limited research dedicated to interventions regarding colorectal cancer surgery and the relatively vast amount of research on colorectal cancer prevention, screening, or diagnosis.^4,5,26,27, 33, 34^ Other colorectal cancer reviews have noted a disproportionately high amount of representation of racial minorities and a very limited representation of the LGBTQI+ community.^30,31^ This is particularly problematic given the colorectal cancer treatment barriers and disparate outcomes noted in the LGBTQI+ population.^32,33,34^

Our systematic review highlights the limited number of studies dedicated to treatment, particularly Surgery. The lack of focus on treatment disparities may provide an insight into the current state of disparities within the United States. For example, current colorectal cancer related death rates are 47% higher in Black men and 34% higher in Black women compared to their White counterparts.^37^ According to our findings, Black patients have been less likely to utilize Surgery for the treatment of colorectal cancer and are twice as likely to refuse Surgery compared to White patients. Further, the laparoscopic approach to resection has been used less often in Black patients (as opposed to the open approach) when compared to White patients. These surgical treatment differences have been found to underlie a substantial portion of the mortality and survival disparities in colorectal cancer outcomes for the Black patient population.^11,14^

### Receipt of Colorectal Surgery

When considering the modern published literature, our systematic review and meta-analysis further suggests that patient medical factors do not fully explain the disparate delivery of surgical care to Black patients.^14^ Additionally, in the context of work by Gill et., al which reported similar odds of receiving colon cancer surgery between Black and White patients in an equal access healthcare system,^38^ it is plausible that access to care is driving some of the observed differences in the utilization of Surgery for colorectal cancer.

### Refusal of Colon Surgery

The limited literature on refusal of surgery also found an increased likelihood of Black patients refusing recommended colon surgery when compared to White patients. Refusal of recommended surgery by Black patients can be attributed to socio-cultural factors including mistrust of the US health care system,^39,40^ the potential for a higher likelihood of poor communication between providers and patients of varying racial backgrounds,^41^ and low health literacy among some Black patients.^42,43^ Interestingly, physicians commonly overestimate patient level of health literacy, especially among Black patients (54%) compared to White patients (11%).^44^ This may be impeding the clarification of common misunderstandings that drive patients to refuse surgery; misunderstandings such as the belief that Surgery enables cancer to spread or confusion regarding the difference between malignant and metastatic disease.^45^

### Laparoscopic versus Open Colorectal Cancer Surgery

Laparoscopic colorectal resection is associated with better overall outcomes, including: lower frequency of blood transfusions and surgical site infections and decreased rates of readmission and mortality.^46,47,48,49^ Additionally, the laparoscopic approach for colon resection has also been shown to result in an 11.3% reduction in postoperative ileus and a shorter length of stay when compared with open surgery.^49^ Our finding that Black patients are less likely to receive laparoscopic surgery, when compared to White patients, may contribute to the observed disparities in surgical outcomes and colorectal cancer outcomes, in general.^50,51^

Patient and provider factors may influence the observed disparities in the receipt of laparoscopic surgery among Black patients. Body mass index (BMI) is the primary factor demonstrated to be a predictor of open versus laparoscopic surgery for colorectal cancer.^52^ It is possible that differences in BMI between the typical Black and White patient may drive the observed differences in receipt of laparoscopic surgery.^53^ If this is the underlying etiology, then consideration for preoperative optimization to overcome this barrier must be pursued.

Studies, that adjusted for BMI, have shown that the underutilization of laparoscopic colorectal surgery cannot be entirely explained by differences in patient characteristics or availability of laparoscopic equipment.^54^ However, geographic and hospital factors are significantly associated with receipt of laparoscopic colorectal surgery, thus potentially influencing a patient’s options for surgical approach.^55,56^ Keller et. al reported that the following factors indicated a higher likelihood of approaching colon cancer laparoscopically: higher volume surgeons (3.5 times), Colorectal versus General Surgeons (1.3 times), and urban versus rural location of hospital (1.5 times). Additionally, hospitals in the Northeast and Western United States were more likely to utilize the laparoscopic approach versus hospitals in the Midwest.^57^ As such, differences in hospital and surgeon selection between Black and White patients may also influence the observed disparity in the use of laparoscopic surgery among Black patients.

### Limitations

This study has several limitations. Based on the existing literature, all of the studies included in the meta-analysis were retrospective. Due to the limitations of the data sources used for the primary studies, longitudinal outcomes could not be evaluated. We observed notable heterogeneity for two analyses: receipt and refusal of colorectal surgery. To address this limitation, we used random effects models for these analyses. For the receipt of surgery analysis, we also conducted separate analyses of nonoverlapping patient groups who received surgery on the colon or the rectum. These individual analyses had limited heterogeneity and the results among all three analyses presented similar odds ratios. Further, studies within the receipt of surgery analysis, are largely limited by the inability to assess whether lack of surgery was due to patient refusal or whether surgery was not presented as an option. The two studies that examined refusal of Surgery had overlapping patient cohorts. Interestingly, the more recent study demonstrated trend towards a greater likelihood for Black patients to refuse surgery than the earlier study. Finally, within the risk of bias assessment for comparability of groups, the primary variable of collecting a measure for comorbidities, was absent in three studies. Two of the studies were used in the receipt of colorectal surgery meta-analysis which may explain the observed high heterogeneity.

## Conclusions

Despite disparities that exist across the continuum of cancer care, the vast majority of research in the past 10 years has remained focused on Prevention, Screening, or Diagnosis within racial minorities. Relatively few studies have addressed disparities within the treatment domain. All of the surgical studies were observational without any studies testing interventions to reduce surgical disparities. Future studies should include an expansion of the existing work to understudied populations such as the LGBTQI+ community. Additionally, because Black patients remain less likely to receive colorectal surgery, twice as likely to refuse surgery, and less likely to receive laparoscopic surgery for colorectal cancer, future efforts to reduce colorectal cancer disparities should include interventions within Surgery

## Data Availability

Online publications
Citations 11-24

## Data Availability Statement

The data that support the findings of this study are openly available in PubMed (https://pubmed.ncbi.nlm.nih.gov/) at

Reference number 11; DOI Link: 10.1016/j.amjsurg.2020.06.020

Reference number 12; DOI Link: 10.1245/s10434-017-6306-4

Reference number 13; DOI Link: 10.1016/j.suronc.2018.11.010

Reference number 14; DOI Link: 10.1158/1055-9965.EPI-20-0950

Reference number 15; DOI Link: 10.1002/cncr.26034

Reference number 16; DOI Link: 10.1007/s00464-017-5782-8

Reference number 17; DOI Link: 10.1002/jso.25917

Reference number 18; DOI Link: 10.1016/s0027-9684(15)30112-7

Reference number 19; DOI Link: 10.1002/cam4.3316

Reference number 20; DOI Link: 10.1097/DCR.0000000000000874

Reference number 21; DOI Link: 10.1097/MCG.0000000000000951

Reference number 22; DOI Link: 10.2105/AJPH.2014.302079

Reference number 23; DOI Link: 10.1097/SLA.0000000000001781

Reference number 24; DOI Link: 10.1002/cncr.32529

## Notes

### Competing Interest Statement

The authors have declared no competing interest.

### Funding Statement

No funding

### Author Declarations

Exempt from IRB review

