## Supplemental File for "Colorectal Cancer Disparities Across the Continuum of Cancer Care: A Systematic Review and Meta-Analysis"

**Table 1.**

**Table 1a. PubMed Original Search Results: Searches limited to publication date January 1, 2011 – March 29, 2021.**

| Search | Query | Results |
|---|---|---|
| #1 | (("colorectal neoplasms"[MeSH Terms] OR ("colorectal"[All Fields] AND "neoplasms"[All Fields]) OR "colorectal neoplasms"[All Fields] OR ("colorectal"[All Fields] AND "cancer"[All Fields]) OR "colorectal cancer"[All Fields]) AND ("disparate"[All Fields] OR "disparately"[All Fields] OR "disparities"[All Fields] OR "disparity"[All Fields])) AND ((humans[Filter]) AND (2011:2021[pdat])) | 1,600 |

**Table 1b. PubMed Additional Search Results: Searches limited to publication date January 1, 2011 – March 29, 2011**

| Search | Query | Results |
|---|---|---|
| #1 | (((Colon Cancer OR Rectal Cancer OR Colorectal Cancer) AND (Race OR Socioeconomic OR Age OR Gender OR Disparities OR Sexuality OR LGBTQIA)) NOT (DNA OR Micro OR Bio OR Hered)) NOT (Stem OR Gene) AND ((y_10[Filter]) AND (booksdocs[Filter]) OR randomizedcontrolledtrial[Filter]) AND (humans[Filter]) AND (English[Filter])) Filters: Books and Documents, Randomized Controlled Trial, in the last 10 years | 816 |

| Table 2. | Characteristics of Included Studies | | | | | |
|---|---|---|---|---|---|---|
| **Study** | **Year** | **Data Source** | **Total Participant Size** | **Primary Disparity** | **Outcome** | **Covariates** |
| Alty IG., et al | 2021 | National Cancer Data Base | 167411 | Race | Refusal of Surgery | Stage, Age, Sex, Education, Annual Income, Insurance, Living Environment, CDCC Score, Year of Diagnosis, Facility Location, Facility Type |
| Arsoniadis EG., et al | 2017 | Nationwide Inpatient Sample | 108437 | Race | Receipt of Surgery *(Rectum)* | Age, Sex, Elixhauser score, and Admission Type, Hospital Factors, Insurance Status |
| Birkett RT., et al | 2019 | Surveillance, Epidemiology and End Results (SEER) Program–Medicare file | 4834 | Race | Receipt of Surgery *(Colon)* | Patient Age, Race, Gender, Klabunde-Charlson Comorbidity Score, Socioeconomic Status, Urban/Rural Location, Tumor Location, Year Of Diagnosis |
| Bliton JN., et al | 2021 | National Cancer Data Base | 219334 | Race | Receipt of Surgery *(Colon)* | Stage, Histology, And Comorbidities And Sex, ZIP Income Quartile And Insurance, And Region, Patient Urban Context, Cancer Center Type |
| Bliton JN., et al | 2021 | National Cancer Data Base | 117897 | Race | Receipt of Surgery *(Rectal)* | Stage, Histology, And Comorbidities And Sex, ZIP Income Quartile And Insurance, And Region, Patient Urban Context, Cancer Center Type |
| Haas JS., et al | 2011 | Surveillance, Epidemiology and End Results (SEER) | 36648 | Race | Receipt of Surgery *(CRC)* | Sex, Age (Continuous), Marital Status, Comorbid Conditions, Year Of Diagnosis, Health Service Area, Urban/Rural, Income, Surgeon Capacity |

| | | | | | | |
|---|---|---|---|---|---|---|
| | | Program–Medicare file, the American Medical Association (AMA) Physician File, and the Area Resource File (ARF). | | | | |
| Hawkins AT., et al | 2018 | National Cancer Data Base | 118735 | Race | Receipt of Laparoscopic vs Open Surgery | Stage, Tumor Size, Patient- And Hospital-Level Demographics, Comorbidities, Insurance |
| Lu PW., et al | 2020 | National Cancer Data Base | 137882 | Race | Refusal of Surgery | Age, Sex, Race, Charlson Comorbidity Score, Clinical Stage, Receipt Of Chemotherapy, Patient Insurance, Patient Income, Facility Characteristics, County Type, Year Of Diagnosis |
| Munene G., et al | 2013 | Nationwide Inpatient Sample | 93542 | Race | Receipt of Surgery *(CRC)* | Patient Demographics, Admission Status, Comorbidities, Hospital Characteristics, And Year Of Admission |
| Raoof M., et al | 2020 | California Cancer Registry | 11255 | Race | Receipt of Surgery *(CRC)* | Facility, Age, Comorbidities, Insurance, Differentiation Of Tumors, Extra-Hepatic Metastases, Marital Status, SES, Side Of Tumor, Peri-Operative Chemotherapy, Type Of Hospital, Hospital Volume, NCI Designation, Hospital Location |
| Ratnapradipa KL., et al | 2017 | National Cancer Institute's Surveillance, Epidemiology, and End Results (SEER) | 9847 | Race | Receipt of Laparoscopic vs Open Surgery | Age, Sex, Race/Ethnicity, Number Of Comorbidities ,Tumor Stage, Grade, Location, Histology, Hospital Size, Hospital Teaching Status, Mean Annual Surgeon Caseload, County-Level Poverty, Rural/Urban |

| | | | | | | |
|---|---|---|---|---|---|---|
| | | database with Medicare claims data (2008–2011), supplemented with county-level demographic data from the 2010 US Census and the 2008–2012 American Community Survey (ACS). | | | | |
| Rodriguez EA., et al | 2018 | Florida Agency for Healthcare Administration Hospital Admission Database | 39767 | Race | Receipt of Surgery *(CRC)* | Smoking, Age, Sex, Charlson-Deyo Comorbidity Score, Insurance Status (Insured Or Uninsured), Presence Of IBD |
| Samuel CA., et al | 2014 | Veteran Affairs Central Cancer Registry | 6548 | Race | Receipt of Surgery *(Colon)* | Age, Gender, Marital Status, And Area-Level Socioeconomic Status, Comorbidities, History Of Any Cancer, Year Of Diagnosis, Tumor Stage, Tumor Grade |
| Samuel CA., et al | 2014 | Veteran Affairs Central Cancer Registry | 1887 | Race | Receipt of Surgery *(Rectum)* | Age, Gender, Marital Status, And Area-Level Socioeconomic Status, Comorbidities, History Of Any Cancer, Year Of Diagnosis, Tumor Stage, Tumor Grade |
| Turner M., et al | 2017 | National Cancer Data Base | 21971 | Race | Receipt of Laparoscopic vs Open Surgery | Age, Sex, Annual Income, Education, Hospital Location And Type, Year Of |

| | | | | | | |
|---|---|---|---|---|---|---|
| | | | | | | Diagnosis, Pathologic Stage, Extent Of Surgery, Insurance |
| Uppal A., et. al | 2019 | National Cancer Data Base | 21155 | Socioeconomic Status (Income Level) | Receipt of Surgery | Age, Ethnicity, Race, Crow Fly Distance, N Classification, Surgical Resection Margins Of The Primary Tumor, Charlson-Deyo Comorbidity Score, Facility Type, FIQ, Use Of Multiple Facilities, Receipt Of Systemic Therapy |
| Uppal A., et. al | 2019 | National Cancer Data Base | 14679 | Race | Receipt of Surgery *(CRC)* | Age, Ethnicity, Race, Crow Fly Distance, N Classification, Surgical Resection Margins Of The Primary Tumor, Charlson-Deyo Comorbidity Score, Facility Type, FIQ, Use Of Multiple Facilities, Receipt Of Systemic Therapy |

| Table 3. | Risk of Bias Assessment | | | | | | | | |
|---|---|---|---|---|---|---|---|---|---|
| Study | Selection | | | | Comparability | Outcome | | | Total Score |
| | Exposed | Unexposed | Exposure | Outcome | | Assessment | Follow-Up | Attrition | |
| Alty IG., et al | * | * | * | * | ** | * | | | 7 |
| Arsoniadis EG., et al | * | * | * | * | * | * | | | 6 |
| Birkett RT., et al | * | * | * | * | ** | * | | | 7 |
| Bliton JN., et al | * | * | * | * | ** | * | | | 7 |
| Haas JS., et al | * | * | * | * | ** | * | | | 7 |
| Hawkins AT., et al | * | * | * | * | ** | * | | | 7 |
| Lu PW., et al | * | * | * | * | ** | * | | | 7 |
| Munene G., et al | * | * | * | * | * | * | | | 6 |
| Raoof M., et al | * | * | * | * | ** | * | | | 7 |
| Ratnapradipa KL., et al | * | * | * | * | ** | * | | | 7 |
| Rodriguez EA., et al | * | * | * | * | * | * | | | 6 |
| Samuel CA., et al | * | * | * | * | ** | * | | | 7 |
| Turner M., et al | * | * | * | * | ** | * | | | 7 |
| Uppal A., et. al | * | * | * | * | ** | * | | | 7 |
